# Feasibility study on a Noninvasive Assessment of ALS Patient Emotional State

**DOI:** 10.64898/2026.03.18.26348710

**Authors:** Marc Garbey, Quentin Lesport, Gülşen Öztosun, Megan Heidebrecht, Kamyab Pirouz, Elham Bayat

## Abstract

This study addresses the need for objective, real-time assessment of emotional responsiveness and coping strategies in individuals with Amyotrophic Lateral Sclerosis (ALS) to support personalized care. We are using non-invasive speech analysis and data science methods on an expanded cohort comprising 28 ALS patient visits.

We first demonstrate that commonly available artificial intelligence tools, including current-generation large language models (LLMs), such as ChatGPT, Gemini and Claude, do not provide reliable or reproducible assessments of patient concern levels in the absence of expert clinical supervision.

Further, we observe a discrepancy between subjective and objective metrics such as the forced vital capacity for breathing.

We introduce a novel functional classification system that contextualizes clinician-rated emotional concern relative to the patient’s functional impairment as measured by the ALS Functional Rating Scale (ALS-FRS). Patient responses are categorized as:

- **Congruent:** Emotional responsiveness is proportional to functional impairment.
- **Muted:** Emotional response is lower than expected given functional impairment.
- **Excessive:** Emotional response exceeds that expected given functional impairment

**Key Findings:** Emotional Drivers and Acoustic Filtering

We subsequently identified objective digital biomarkers, independent of LLMs, that robustly distinguish the muted and excessive response groups. Our data analysis revealed that the excessive response group was predominantly male and required significantly more clinician time, suggesting that this type of classification is primarily driven by psychosocial coping mechanisms such as anxiety related to functional dependency. Conversely, the muted response group was predominantly female.

Most significantly, we discovered a direct link between this behavioral classification and the patient’s speech acoustics, demonstrating that dysarthria may act as an acoustic filter for emotional expression:

1. Patients with muted response exhibited a voice profile characterized by high acoustic loudness, high sharpness, low roughness, and low fluctuation consistent with attenuation of emotional expression.
2. Patients with excessive response presented with the opposing profile: low loudness, low sharpness, high roughness, and high fluctuation consistent with amplification of emotional expression.

However, a significant diagnostic overlap exists between these biomarkers of anxiety and the early manifestations of Amyotrophic Lateral Sclerosis, spastic dysarthria for the former, flaccid dysarthria for the second.

Our work enables non-invasive assessment of both emotional coping strategies and the dominant underlying neuromuscular mechanisms. These insights support the early identification of patients who may benefit from targeted, proactive interventions, and represent a significant step toward personalized ALS management.

## Introduction

Amyotrophic lateral sclerosis (ALS) is a progressive neurodegenerative disease that severely impairs a patient’s ability to communicate through verbal means, facial expressions, and body language, making patient care particularly challenging for healthcare professionals [1]. To maximize information gained during brief clinic visits and support positive adaptation, the multi-disciplinary team must accurately appreciate the patient’s emotional responses [2], [3]. However, while psychological states vary greatly and may be better than expected, detailed psychological assessments are often not part of routine visits [4], and even experienced physicians find interpreting non-verbal communication of emotions difficult. Consequently, clinicians must often discuss critical treatment choices—such as tube feeding and respiratory support—and the use of invasive supportive interventions with patients who are frequently unprepared. Maintaining patient morale is essential for effective patient-physician collaboration, requiring the physician to accurately assess the patient’s emotional state and balance their subjective experiences with objective medical conditions. Failure to anticipate or understand potential emotional disconnections (e.g., denial or overreaction) can severely undermine the therapeutic relationship [5].

Recognizing this critical clinical need, our previous work addressed the challenge of assessing emotional state in ALS, which compromises the standard metrics relied upon by traditional affective computing approaches [6], [7], [8]. We developed and validated a specialized, multimodal methodology for interpreting the nonverbal emotional state of early-stage ALS patients [9]. We circumvented muscle weakness—which impairs clear facial expression, speech, and body language [10], [11], [12]—by utilizing physiological signals (pulse rate variability) to quantify stress, combined with Natural Language Processing (NLP) analysis of patient speech and text. This study demonstrated the feasibility of integrating these stress and communication annotations within a Large Language Model (LLM) framework to produce an Automated Patient Priorities Report. This novel report objectively synthesizes the patient’s emotional state, communication content, and stress levels to establish and highlight their immediate concerns, thereby laying a validated foundation for integrating AI tools to generate actionable, personalized clinical insights.

In this follow-up paper, we return to the fundamental clinical problem: the necessity of understanding and anticipating emotional disconnects (such as denial or overreaction) to deliver patient-specific communication strategies that enhance care. We first demonstrate that the unsupervised use of LLMs alone to analyze the emotional state of these patients does not provide a satisfactory, clinically relevant solution. Crucially, this work introduces a novel, clinically-derived classification system based on the congruence between a patient’s emotional response and their objective functional impairment, as measured by the ALS Functional Rating Scale (ALS-FRS). Patients are categorized into three distinct groups:

1. Congruent: Emotional response aligns with functional impairment.
2. Muted: Response is less than expected given the ALS-FRS score.
3. Excessive: Response is greater than expected given the ALS-FRS score.

Furthermore, we identify digital biomarkers—independent of LLMs—that specifically characterize the muted and excessive response groups. Since these two patient categories are at higher risk and require specialized, proactive care, this research marks a significant step toward truly personalized ALS management.

## Methods

### 2.1 Human Subjects

Twenty-five subjects were recruited from the multidisciplinary ALS clinic at George Washington University (GWU). Eligibility required clinical and electrophysiological confirmation of Amyotrophic Lateral Sclerosis (ALS), age over 18 years, and the ability to provide written informed consent. Exclusion criteria included: an unstable medical condition that posed a risk to participation (per the examining physician’s judgment), pre-diagnosis cognitive impairment, dementia, cardiovascular conditions related to arrythmias or a psychiatric illness. Patients with late-stage disease necessitating tube feeding, inability to walk, or significant dysarthria were also excluded. All patients had to be able to talk in an intelligible way, so the disease is early stage with respect to that skill. The study protocol was approved by the GWU Institutional Review Board. The recruitment was done in two stages. the first cohort of 14 patients as described in our initial pilot study [9] that was used to get our first results on ALS patient management and a year later a second cohort to start the process of a longitudinal study published eventually later. This one-year interval between the two cohorts allows us to consider that the first and second visit of the three ALS patients involved in both cohorts will be considered as independent data points for the purpose of this study.

### 2.2 Study Protocol

Patients completed the ALS Functional Rating Scale (ALS-FRS) prior to the clinical examination [13]. Subjects participated in the study during their routine clinic visit with no prolongation of the typical appointment time. Patients’ voices were recorded during the entire visit. Three out of 25 patients had two visits separated by a time lapse over a year. The data set we used for our analysis was 28 visits considered as independent events.

Subjects were asked to respond to four questions by the neurologist to assess emotional response (Table 1). If the patients were already enrolled in the previous pilot study and had any interventions since then, modified versions of the same questions were asked accordingly.

**Table 1.**
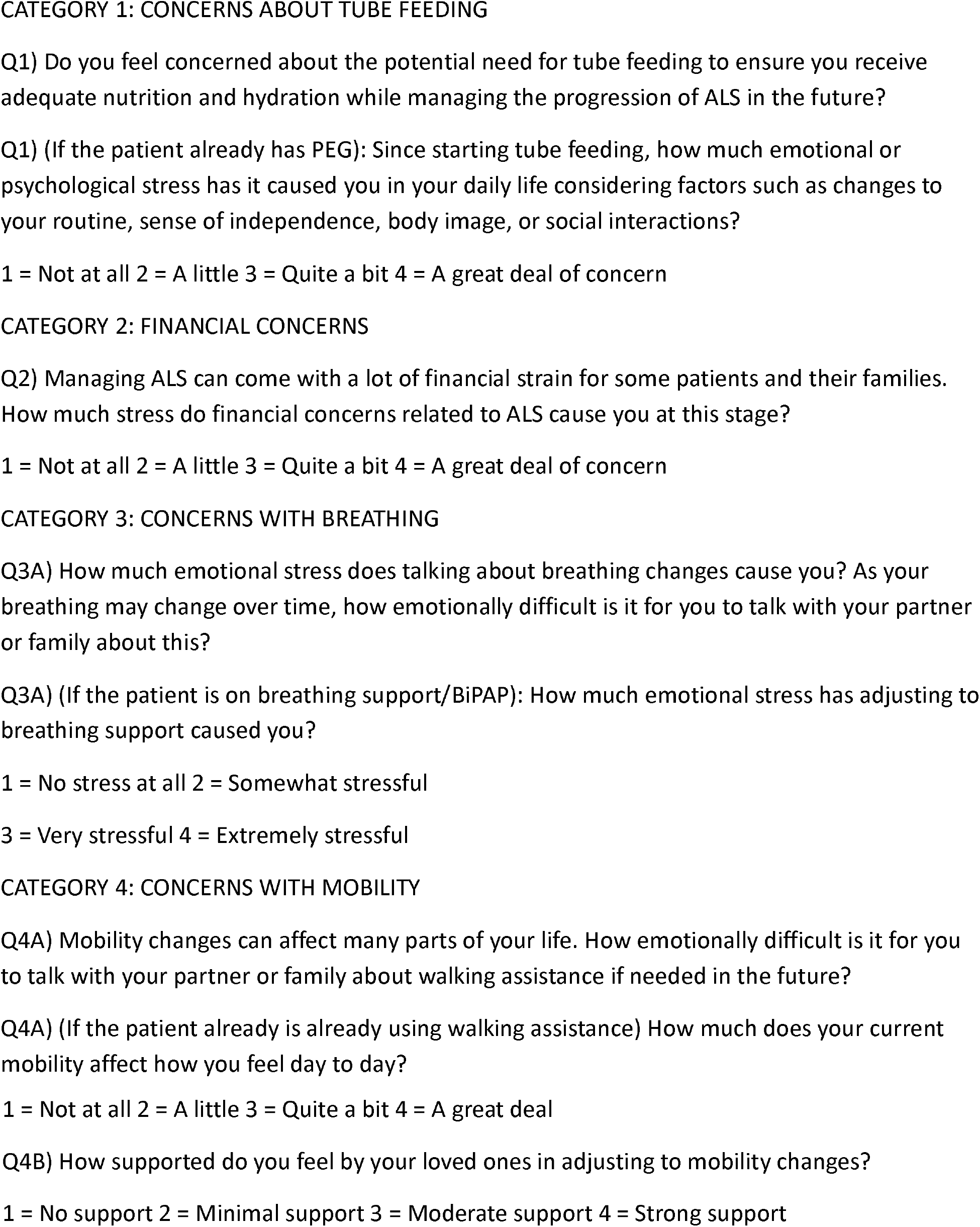
Emotional Response Questions in 4 Categories and Their Ratings.

The questions were asked separately and integrated in the conversation. To maintain a natural flow in the conversation, the exact wording varied slightly, while the intended meaning remained the same. The score was a combination of patient feedback and clinical judgment, as it came out from the conversation. Consequently, this process differed substantially from having the patient complete a form independently.

A high-fidelity Zoom-H1n audio recorder (Zoom Corp, NY) captured audio during the clinical session only. This recorder features stereo X/Y 90° microphones, handles up to 120 dB SPL and supports various audio formats.

Building on insights gained from the first cohort’s data collection [9], we introduce in the second cohort a more extensive protocol as follows.

To address the potential impact of anxiety and depression as well as mild cognitive impairment and frontotemporal dementia (FTD) on emotional arousal during the clinical visit we collected the following standard test:

- the Hospital Anxiety and Depression Scale (HADS) [14].
- the Montreal Cognitive Assessment (MoCA) version 7.3 that is appropriate for mild cognitive impairment [15].

To compare the subjective feedback of the patient during the clinical examination with unbiased clinical assessment of physical condition, we collected the following additional tests that are part of standard of care:

- For breathing: Forced Vital Capacity (FVC):
- For Muscle strength: Grip strength in pounds (lbs.) as unit via Hand-Held Dynamometer (HHD)

### 2.3 Natural Language Processing

We leveraged an AI transcription tool, AssemblyAI - https://www.assemblyai.com/-, to generate transcripts of the recorded clinical sessions. These transcripts included timestamps for each word, confidence scores, and speaker identification. The transcripts were segmented into sentences categorized by speaker (doctor, patient, or companion) and separated questions from statements. To facilitate topic classification, all transcripts were anonymized and combined into a single file. We then built a dictionary of key terms for classifying sentences into six categories related to nutrition, breathing, walking, speech, medication, finance, and miscellaneous. This dictionary development employed a combination of manual extraction and generative language processing using ChatGPT. The dictionary has the capability to be refined and enhanced to minimize NLP misclassifications. Our analysis yielded several key metrics, including the relative weight of conversation topics and subject participation levels.

### 2.4 Large language Model

We ask each of these three LLMs, namely ChatGPT from Open AI -https://openai.com/, Gemini from Google and Claude from Anthropic, to evaluate the level of concern of patients with respect to Q1 to Q4 as in table 1.

The prompt was stated as: “From the following conversation, can you score successively from 0 to 3 the anxiety of the patient related to 1. breathing issues, 2 Mobility issues, 3 Nutrition issues, 4 Finance issues. 0 meaning no anxiety, 1 meaning mild anxiety, 2 meaning moderate anxiety and 3 meaning extreme anxiety”.

The conversation that was provided was the AI transcription annotated by who is speaking, i.e. the clinician, patient or companion if any, to avoid any confusion.

The results were archived in an excel file and could be compared to the evaluation of the experienced neurologist who ran the clinical examination.

To assess the reproducibility of the LLM answers we submitted the same prompts to each LLM in separate conversation separated by few days. The intent was to run the same version of the LLM to make a fair comparison. As a matter of facts every few months, respectively weeks, one of these LLM version – respectively subversion-would be released. The consistency and fidelity of the answer between two same prompts should be of prime concern as patient may not appreciate a that answers may have such a degree of randomness.

As opposed to our previous pilot study [9] the clinical conversation was submitted to the LLM as is, i.e. without any preprocessing: we did not use a subset of the full transcript containing key moments identified through affective computing methods described in [10] as we hypothesized that this condensed transcript captures the most relevant aspects of the clinical conversation.

### 2.4 Metrics and Classification

To investigate the relationship between various human factors and AI analysis results, we performed a combination of correlation and classification analysis. The correlation study assessed metrics including but not limited to sentiment analysis scores, clinical examination duration, patient self-assessment feedback and scores of standardized clinical assessments such as HADS, MoCA and ALS-FRS. As mentioned earlier, the HADS and MoCA assessments were part of the second cohort, not the first one: therefore, our sample size was relatively small, i.e. 14 visits. For clarity, each correlation result is presented as a triplet: the Pearson coefficient (strength and direction), the P-value (statistical significance), and the sample size (denoted N). We also included scatter plot visualizations to graphically represent the relationship and overall trend. It is important to note that while these metrics offer valuable insights, they do not establish causality.

As we construct the following classification system based on the congruence between a patient’s emotional response and their score on the ALS Functional Rating Scale (ALS-FRS):

1. Congruent: Emotional response aligns with functional impairment.
2. Muted: Response is less than expected given the ALS-FRS score.
3. Excessive: Response is greater than expected given the ALS-FRS score.

We need to establish if the corresponding feature data sets of this classification are truly different. We perform the Wilcoxon Rank sum test, observing that all three sets do not have the same median. To that end, we use the one side ranksum matlab function. The Wilcoxon Rank-Sum Test is a robust non-parametric test that can be used to compare two small independent groups. Unlike the t-test, it does not assume the data follows a normal distribution.

### 2.5 Affective Computing

We focus on a non-intrusive data acquisition that uses exclusively the speech of the patient. To assess patient emotional arousal, we employed the following techniques:

Sentiment Analysis: We used the Valence Aware Dictionary and Sentiment Reasoner (VADER) sentiment lexicon [16] to analyze the transcript text and identify sections with the most negative sentiment, potentially reflecting emotional distress. This sentiment analysis approach can be further refined and specialized to ALS patients as a larger data set is analyzed.

#### Voice Analysis

From the speech sound signal we compute the following 9 key Psycho-acoustic features [17]:

- The acoustic loudness according to ISO 532-1 (Zwicker) or ISO 532-2 (Moore-Glasberg).
- The acoustic Sharpness (Sharpness relates to the spectral content, with higher frequencies contributing to higher sharpness).
- Mean, median and maximum values of speech roughness. Roughness is related to rapid amplitude and frequency modulations.
- Mean, median and maximum values of speech fluctuation. Fluctuation is related to slower amplitude and frequency modulations.
- Pitch – the estimates of the fundamental frequency over time for the audio signal.

The relationship between psychoacoustic features and anxiety stems from involuntary physiological arousal that overrides conscious vocal control [18]. In an anxious state, sympathetic nervous system activation increases laryngeal muscle tonus [19], which sharpens the fundamental frequency and restricts natural pitch variability—a phenomenon consistently identified as a primary indicator of stress [20]. Furthermore, the cognitive load associated with anxiety disrupts motor planning, leading to an increased frequency of filled pauses and hesitations [18]. Because these psychoacoustic shifts occur at a level of neuromuscular coordination that is nearly impossible to mask, they serve as reliable, objective markers of a speaker’s internal state, often revealing anxiety even when the speaker attempts to maintain a calm linguistic expression [18], [19].

However, a significant diagnostic overlap exists between these biomarkers of anxiety and the early manifestations of Amyotrophic Lateral Sclerosis (ALS), which alters speech through progressive muscular and respiratory decline [21]. While both conditions may elevate pitch or destabilize voice quality, the underlying drivers differ: anxiety reflects transient autonomic arousal, whereas ALS reflects persistent neurogenic spasticity or weakness. To address this, our framework moves beyond absolute acoustic values, which may be confounded by ALS pathology. Instead, we utilize the relative variation of these biomarkers within the early-stage ALS population to classify patients into the three distinct classification profiles we introduced above: “normal,” “muted,” or “excessive” emotional responses to their symptoms [22], [23]. This comparative approach improves the isolation of psychogenic fluctuations from the pathological baseline of neurodegeneration.

## Result

### Patient Demographics and Baseline Functional Status

We first characterize the study population. The age distribution is presented in Figure 1 (top). The study sample comprises 13 men and 12 women, suggesting that patient recruitment was not significantly biased by sex.

**Figure 1.**
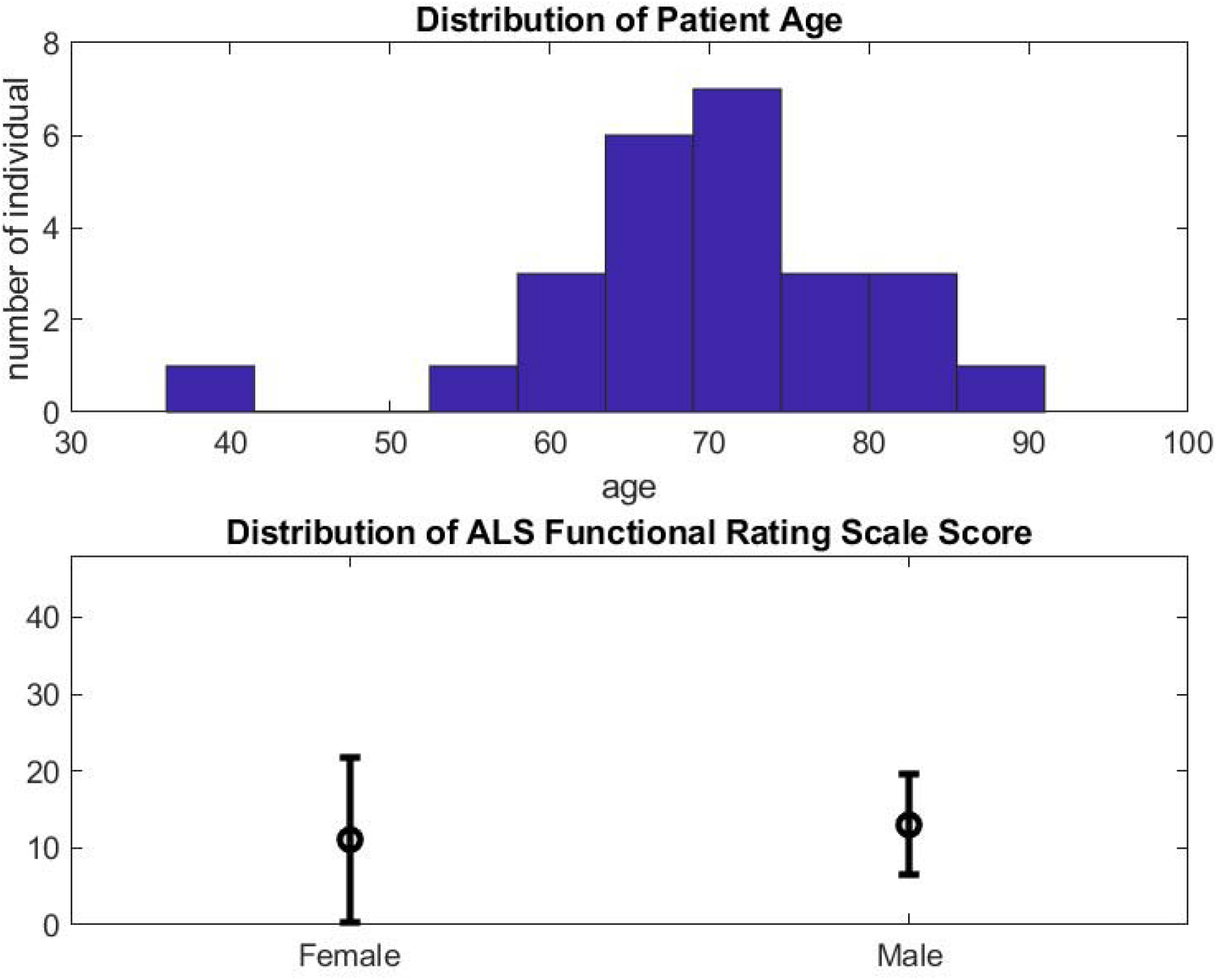
Characteristics of the Population of 25 patients, including 13 males and 12 females: Top: distribution of patient age, bottom: average and standard deviation of the ALS functional rating score for female (left) and male (right).

The global functional score, calculated as the sum of scores from the full functional evaluation (ALS-FRS), is compared between females and males in Figure 1 (bottom). We observe that these two groups exhibit similar global functional scores. The functional assessment, illustrated by the heat map in Figure 2, indicates that all patients were in a relatively early stage of ALS at the time of the clinical visit.

**Figure 2.**
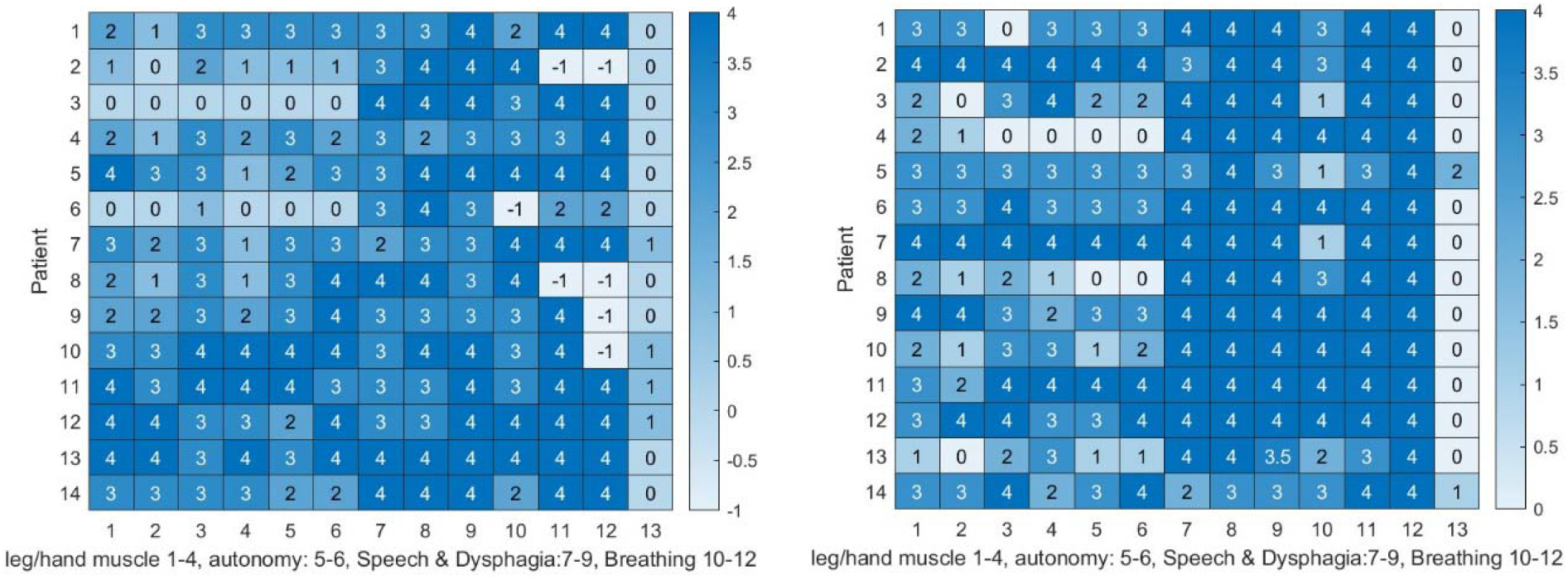
Heatmap summarizing the ALS-FRS scores for all subjects, visually representing the disease severity and progression, with scores (4 = normal, 0 = the worst, -1=N/A). The order of entries corresponds to the ALSFRS categories: leg muscle weakness minutes, hand muscle weakness, autonomy (dressing, hygiene, turning in bed), speech/salivation/dysphagia, and breathing (dyspnea, orthopnea, difficulty breathing). The last column is 0 for spinal ALS onset and 1 for bulbar onset. Functional Assessment of ALS Patients in both cohorts Illustrated by a Left figure corresponds to the first cohort visit, right figure to the second cohort visit

### Psychometric and Cognitive Assessment

In the second cohort of our clinical study, consisting of 14 visits, we introduced the HADS) and MoCA to establish baseline measures of emotional and cognitive health.

The HADS score is interpreted as follows: a score of 0–7 indicates a normal level of anxiety/depression; 8–10 suggests a marginal or borderline abnormal presence of a mood disorder; and 11–21 corresponds to a strong probable presence of a clinically significant mood disorder.

- Figure 4 (right) uses two vertical and horizontal lines to separate patients with normal mood from those with marginal or severe symptoms in the two-dimensional anxiety-depression space.
- The results, shown in Figure 4 (right), indicate that most patients did not have severe signs of depression or anxiety.
- Only 3 out of 14 patients presented with marginal symptoms of depression. 3, respectively 2, had marginal, respectively severe symptoms of anxiety.

We observe a trend in Figure 3 suggesting that sex may influence anxiety scores, with females tending towards higher scores, while the functional score remains comparable for both sexes, as noted previously. A one-sided Wilcoxon Rank Sum Test, assuming the female result median is larger than the male median, yielded p-values of 0.21 for anxiety. Thus, the null hypothesis (no difference in medians) cannot be rejected at conventional significance levels.

**Figure 3.**
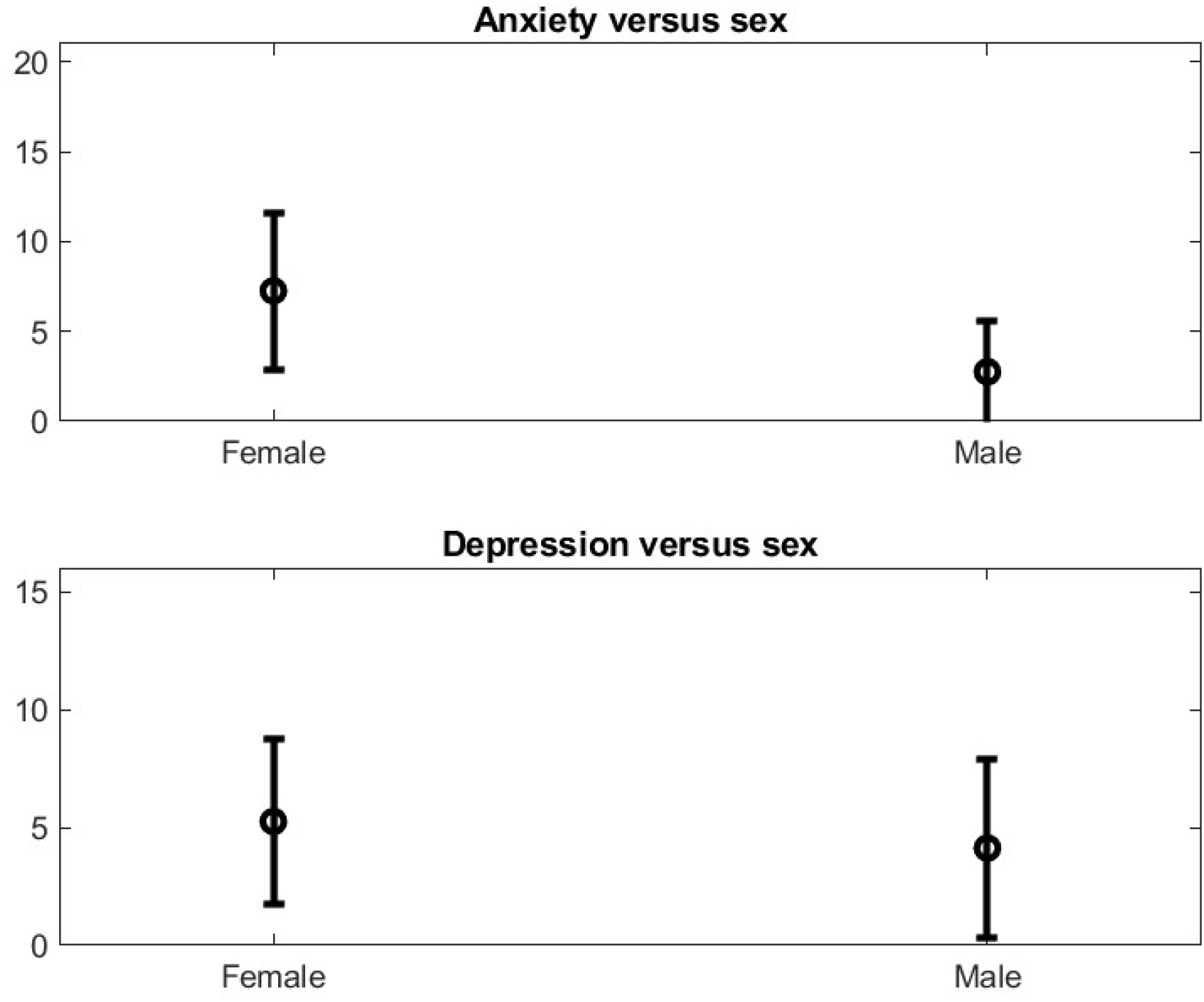
Mean and standard deviation of the anxiety score (top figure) and depression (bottom figure) for the subgroup of males and females using the HAIDS scale. We observe a weak Bias on anxiety (top figure) for Female versus Male on HADS while both subgroups have about the same ALS functional score average.

Regarding the MoCA test, all 14 patients in the second cohort demonstrated normal cognitive function, except for two who showed mild cognitive impairment with respectively MOCA score of 18 and 19 during the first visit. Notably, both of these patients reported very low levels of anxiety. Additionally, 2 of the patients had a modified MoCA score (out of 25 instead of 30) due to hand weakness and could not complete the first section of the assessment, hence lower scores. And another patient did not attempt MoCA.

### Discrepancy Between Subjective and Objective Metrics

We anticipated that subjective feedback on symptoms from patients might differ from objective lab metrics. A notable example is the relationship between FVC, an objective measure of respiratory function, and the patient-reported symptoms of respiratory insufficiency and orthopnea on the ALS-FRS score.

- Figure 4 (left), reporting on the 14 visits of the second cohort, illustrates this discrepancy.
- Some patients with severe breathing distress (low FVC) do not report a particularly bad score for respiratory insufficiency and orthopnea on the ALS-FRS.
- Conversely, a patient with FVC close to 100% may still report severe orthopnea issues.

**Figure 4.**
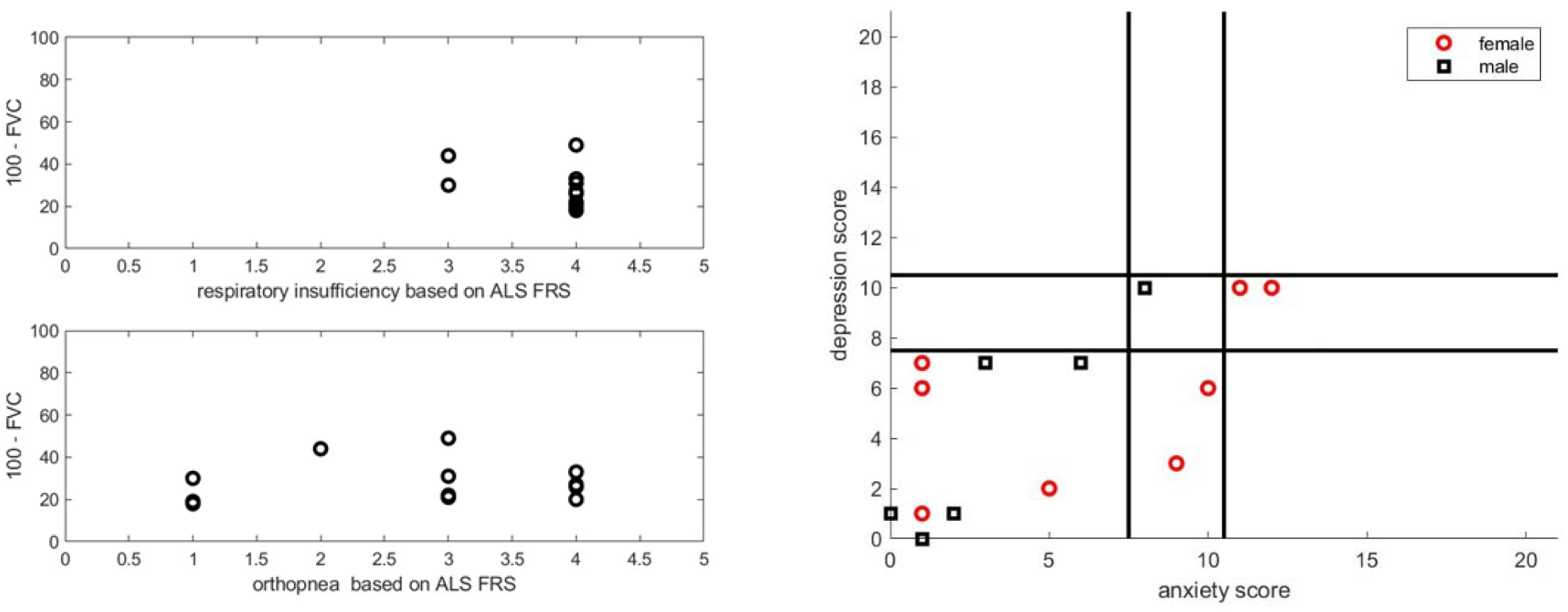
Left figure: on top the Lab Measure of FVC in relation to Patient ALS FRS score patient feedback for breathing. We observe that there is no correlation between the objective metric and subjective metric for the same respiratory function. The vertical axis shows the difference 100-FVC, i.e. the smaller is the number, the better is the breathing capacity. As expected the orthopnea reported symptoms has no correlation with FVC either. Right figure: two-dimensional Representation of the Population HADS Distribution in the anxiety – depression space. The higher the score the worst is the patient mood. The horizontal and vertical lines separate the patient into normal score, marginal score and severe score.

Similarly, the ability for the patient to cut and handle food according to the ALS-FRS is loosely related to the grip strength measured via HHD.

### Evaluation of Large Language Models (LLMs) for Emotional Assessment

The results on the quantitative metric of quality by comparing LLM output to the clinician’s score on four central quality-of-life questions (nutrition, mobility, breathing, and finance burden) were as follows:

- Currently, the LLMs tested were incapable of reproducing the clinician-rated anxiety score with acceptable reproducibility (Figure 5). Using two runs of the same transcript per LLM at some time of interval, we found that 40%, respectively 80% and 20%, of the output shows a maximum difference in question score larger than 2 respectively for Chat GPT, Gemini and Claude.
- Crucially, the uncertainty on the global score—derived from two runs of the same transcript per LLM—is approximately the same order as the difference between the LLM score and the clinician score. This difference is about 2 in the Euclidean norm of the four-dimensional vector difference of scores (maximum score difference is 4).
- In fact, as shown in Figure 2A the LLM gives more diffuse distribution results on question score than the clinician.
- The failure of the LLMs to consistently replicate their own results, illustrated by the high uncertainty, leads to the conclusion that unsupervised LLMs should not be used to write a trusted emotional report on ALS patients (Figure 5).

**Figure 5.**
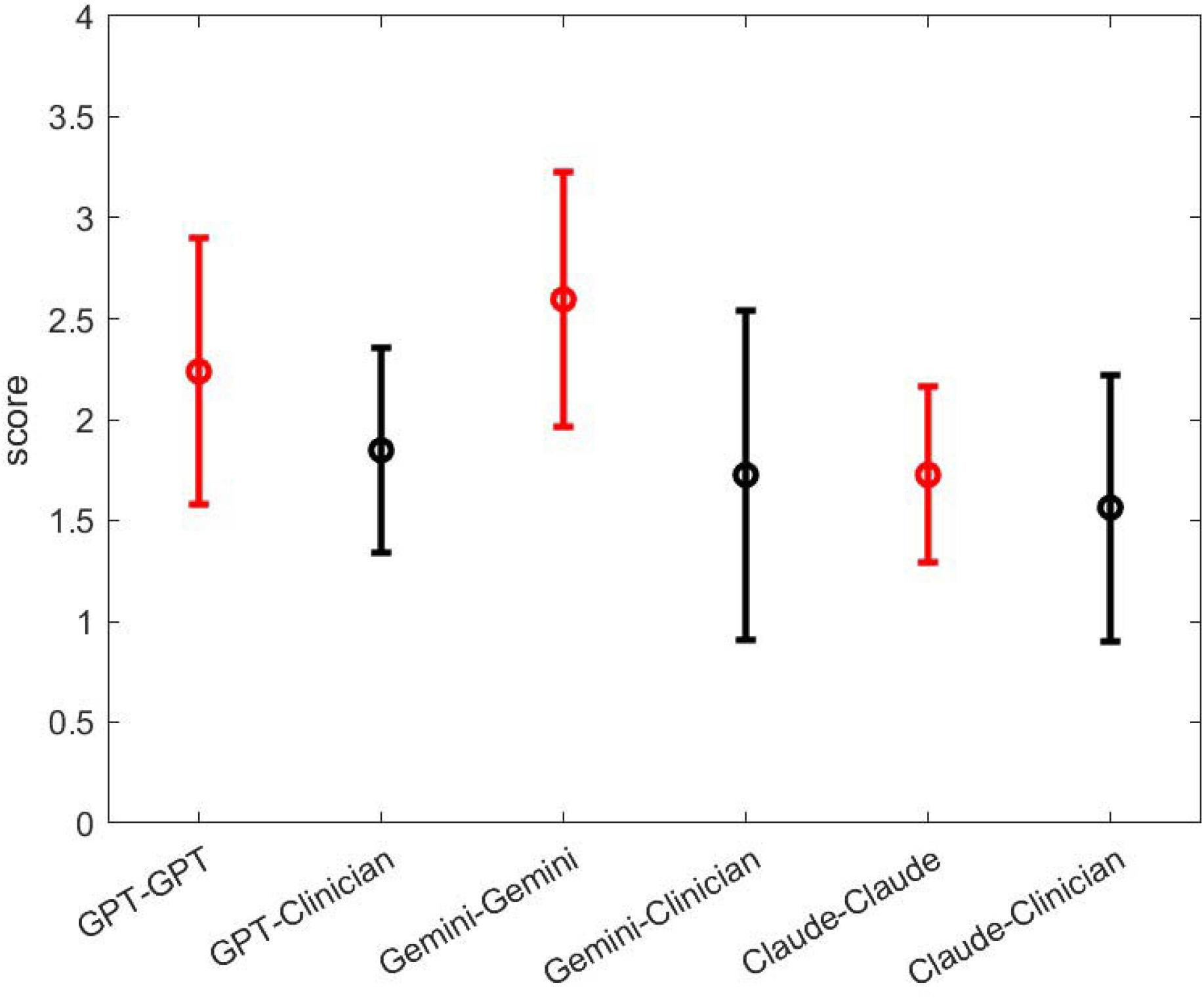
Quantitative assessment of LLM Precision on breathing, mobility, nutrition and finance patient issues with cohort 1. This graphic represents the mean and standard deviation of the difference between two prompt (red bar) and the average of both LLM runs with respect ot the clinical evaluation. For this purpose, we have sum-up the score for each question Q1 to Q4: the minimum score is one and the maximum score is then 16.

We note that Claude from Anthropic yielded significantly better results than ChatGPT and Gemini, suggesting that models specifically tuned for human-like conversation may perform better in this context.

### Patient Classification Based on Subjective Metrics

To assess the congruence between self-reported disease progression (ALS-FRS) and the clinician-rated emotional reaction (anxiety score), we classified patients based on the linear relationship between these two subjective metrics. Figure 6 plots the global anxiety score (sum of the 4 anxiety questions) versus the global ALS-FRS score.

**Figure 6.**
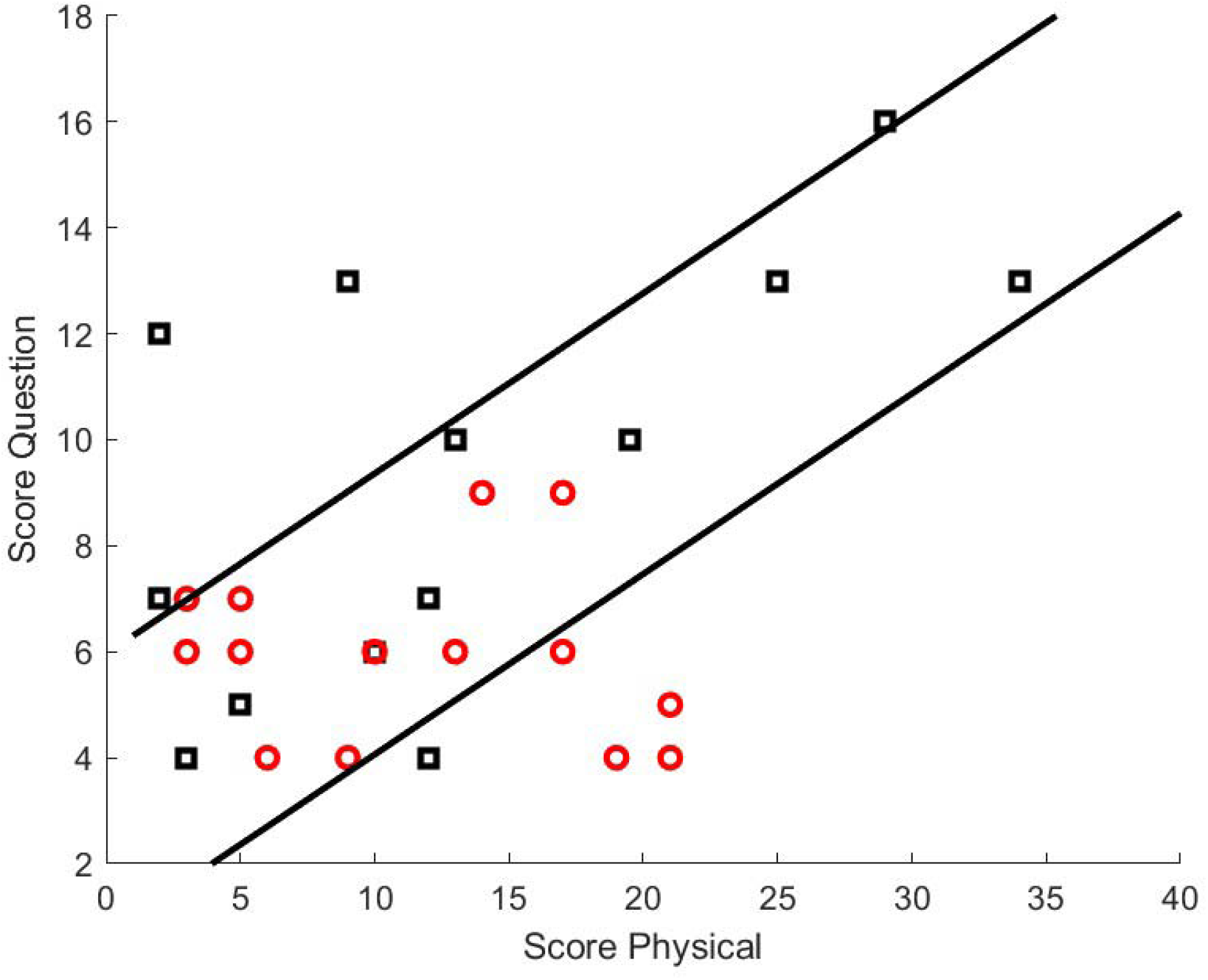
total score on clinical evaluated question Q1 to Q4, versus the total physical functional score with ALS. The diagonal lines separate the patients into three classes: congruent reaction inside the diagonal strip, overexpress response above the diagonal strip (4 patients), muted response below the diagonal strip (5 patients). The lower the physical functional score the better the patient feels, The higher the clinical question score the more the patient is concerned Black square are representing male, red circle are representing female. The correlation coefficient of the question score versus physical score for the set of patient with congruent answer is 0.86, which satisfies our hypothesis that normal reaction would be such.

We assumed that the ALS-FRS score should be linearly correlated with the clinician’s anxiety score for individuals with “congruent behavior”. All three classes are separated as shown in Figure 6. Patients with “muted” response, respectively “excessive” response are below the refined strip, respectively above the defined strip. Patients with congruent response are within the defined strip.

The classification boundaries are defined by the linear equations 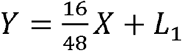 and 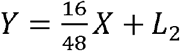.

For Figure 6, the parameters were set to *L*_1_ = 0.5 and *L*_2_ = 6. This setting yielded a significant linear correlation coefficient ρ = 0.86 with a p-value of *p* = 2 × 10^−6^ for the “congruent” response strip.

The distribution of patients across the classes is: 5 in Class 1 (“Muted”), 19 in Class 2 (“Congruent”), and 4 in Class 3 (“Excessive”). This high proportion of congruent responses is consistent with the HADS feedback, where most patients reported normal mood levels. A sensitivity analysis confirmed that the chosen parameters (*L*_1_ = 0.5 and *L*_2_ = 6) represent a suitable compromise, allowing for enough individuals in the extreme classes for feature analysis while maintaining a significant correlation for the congruent class.

### Characteristics of Patient Classes

We next focused on the features that characterize the “Muted” (Class 1) and “Excessive” (Class 3) groups versus the “Congruent” (Class 2) group (Figures 7 & 8).

**Figure 7.**
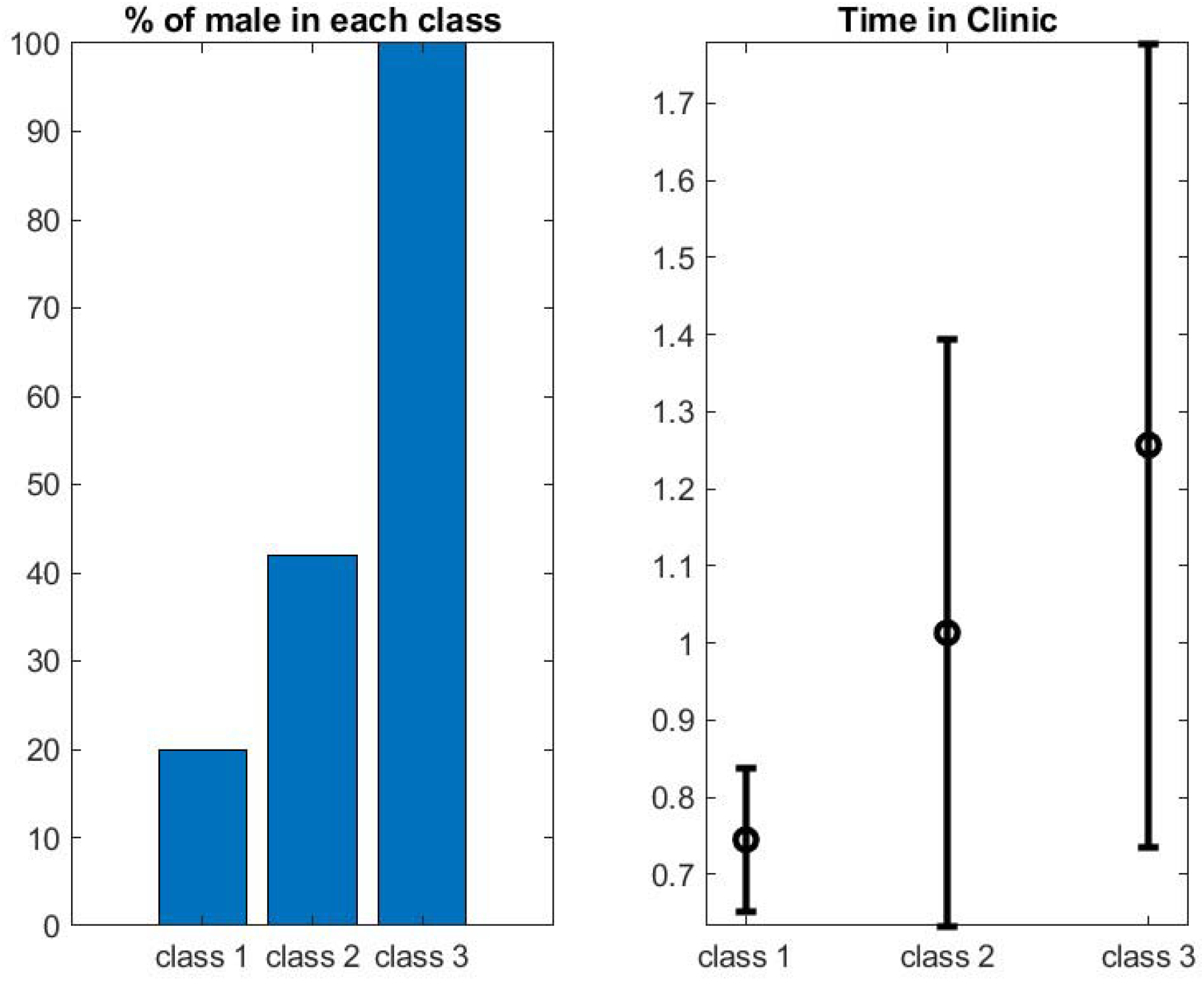
Left figure shows the distribution of sex per class: Class 1 (muted response) has one male and 4 females, and class 3 (excessive reaction) has 5 males and no female. Right figure shows the corresponding average and standard deviation of the time spend in clinic by the patient.

**Figure 8.**
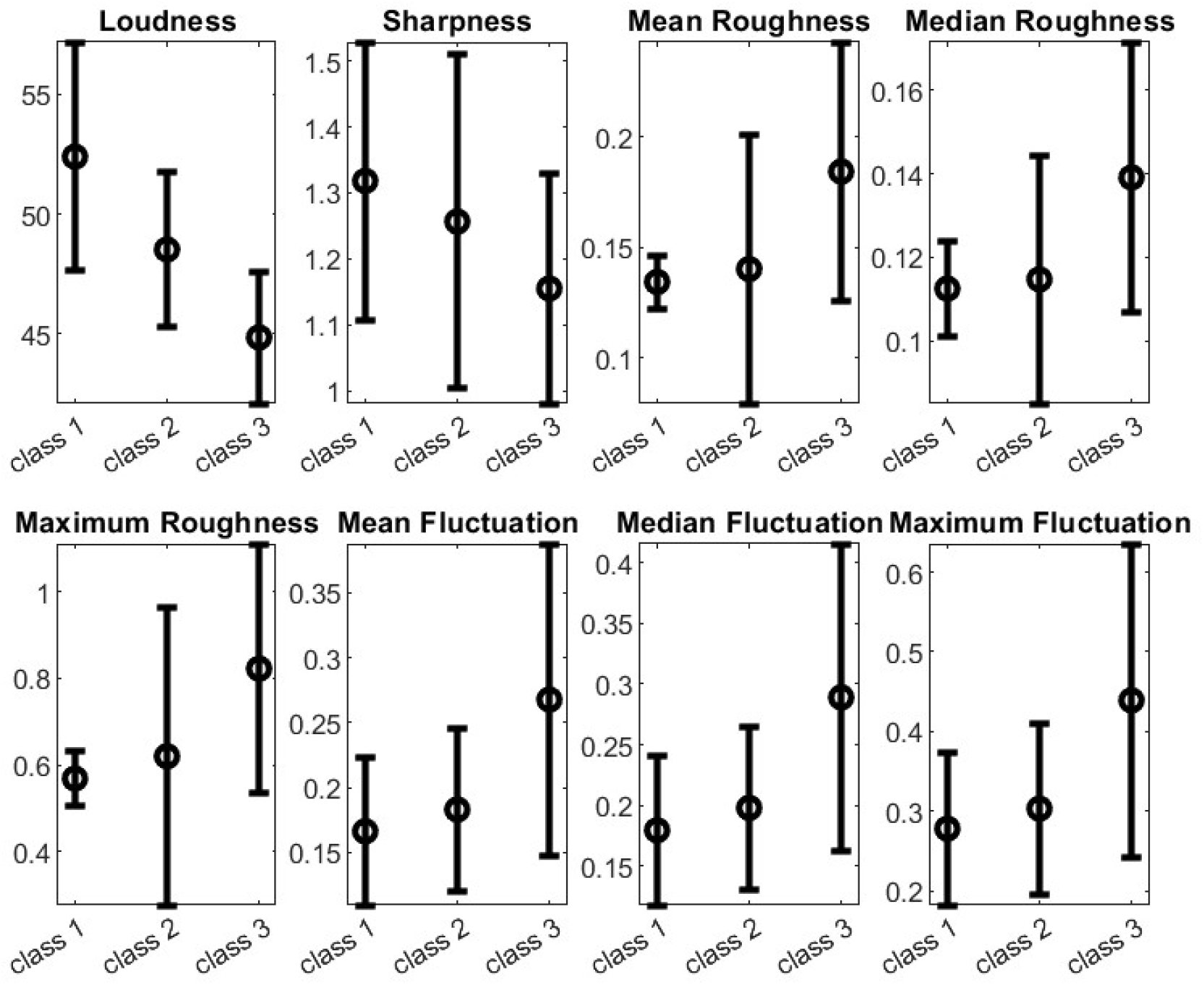
Pattern on class feature mean and std obtained across 8 key Psycho-acoustic features: - The acoustic loudness according to ISO 532-1 (Zwicker) or ISO 532-2 (Moore-Glasberg).
- The acoustic Sharpness (Sharpness relates to the spectral content, with higher frequencies contributing to higher sharpness).
- Mean, median and maximum values of speech roughness. Roughness is related to rapid amplitude and frequency modulations.
- Mean, median and maximum values of speech fluctuation. Fluctuation is related to slower amplitude and frequency modulations. Each bar represents the average and standard deviation of the digital metric per class of patient.

Two clear differentiators between the three classes are the percentage of males and the time spent in the clinic. The longer time spent in the clinic for the “Excessive” group is consistent with expectations. The finding regarding the percentage of males and female in each class will be discussed further. We found no significant quantitative differences for the three classes regarding the conversation time distribution of topics or the percentage of the session talked by the patient. Sentiment analysis also did not provide a clear differentiator.

A key noninvasive finding is based on the psycho-acoustic characteristics of the patient’s speech. Differences in acoustic loudness and roughness between the “muted” and “excessive” classes are highly significant (one-sided Wilcoxon Rank Sum Test, Table 2). Acoustic features such as fluctuation (or tremor), pitch, and sharpness provided a more nuanced result, showing a monotonic trend for sharpness and fluctuation with respect to our classification (Figure 8), though the one-sided Wilcoxon Rank Sum test did not yield a definitive conclusion ρ ≈ 0.15)

**Table 2:** p-value for the Wilcoxon rank sum test – left and right corresponds respectively to the assumption of the median of the first set is less, respectively more than the median of the second set.

|  | Time in Clinic | Loudness | Sharpness | Mean Roughness | Median Roughness | Max Roughness | Mean Fluctuation | Median Fluctuation | Max Fluctuation |
| --- | --- | --- | --- | --- | --- | --- | --- | --- | --- |
| Classes 1&2 | 0.03 left | 0.08 right | 0.33 right | 0.21 left | 0.5 left | 0.19 left | 0.36 left | 0.33 left | 0.41 left |
| Classes 2&3 | 0.2 left | 0.05 right | 0.27 right | 0.03 left | 0.07 left | 0.02 left | 0.09 left | 0.08 left | 0.09 left |
| Classes 1#3 | 0.016 left | 0.016 right | 0.09 right | 0.06 left | 0.09 left | 0.03 left | 0.09 left | 0.09 left | 0.14 left |

These vocal features suggest the acoustic capture of the two dominant subtypes [24], [25], [26]:

1. “Muted Response” (Class 1): Patients exhibited a voice profile characterized by high acoustic loudness, high sharpness, low roughness, and low fluctuation. This acoustic signature—a tense, regular, and bright voice—possibly expressing an excessive laryngeal hypertonicity, often described as a “strained-strangled” vocal quality.
2. “Excessive Response” (Class 3): Patients presented with the opposing profile: low loudness, low sharpness, high roughness, and high fluctuation. This acoustic pattern—a weak, dull, and highly unstable voice is leading possibly to a hoarse and highly unstable vocal output.

Finally, as pitch is a core prosodic feature [27], we found that sentiment analysis and pitch evaluation in our population sample are significantly correlated with ρ = 0.74 and a p-value of 6.4 × 10^−6^ (Figure A1).

A sensitivity analysis on the parameters *L*_1_ and *L*_2_ was performed:

- As *L*_1_ approaches *L*_2_, the “congruent” strip narrows, causing the feature differences between Class 1/3 and Class 2 to diminish.
- Conversely, for a wider strip (smaller *L*_1_ and larger *L*_2_ ) the extreme classes (1 and 3) contain fewer individuals but show about the same feature differences.
- The chosen values of *L*_1_ =0.5 and *L*_2_=6 were determined to be a good compromise, ensuring sufficient individuals in the extreme classes for robust feature analysis while maintaining a significant correlation.

We will discuss these findings further in the next section, in particular a novel objective link between behavioral response patterns and underlying neuropathology as a significant diagnostic overlap exists between these psychoacoustic biomarkers of anxiety and the early manifestations of Amyotrophic Lateral Sclerosis (ALS).

## Discussion

The non-invasive assessment of a patient’s emotional state, particularly in a complex, progressive condition like ALS, is a notoriously difficult problem [10], [28], [29]. Our study introduces and validates a novel functional classification approach, demonstrating that observed behavioral patterns of emotional expression are deeply interconnected with both psychosocial coping mechanisms and underlying neurological pathology.

In this manuscript, we adopted a pragmatic criterion: evaluating whether a patient’s level of concern is proportional to the nature of their physical symptoms (behaving “congruently”), or whether they exhibit an “excessive” or “muted” reaction. By focusing on emotional management rather than intrinsic emotional state, we shift the clinical focus to how interventions can best support patients coping with ALS progression. While it is tempting to link excessive reaction to anxiety and muted reaction to depression, we acknowledge this is a simplification and have not rigorously established such a psychological correlation in this study.

While this feasibility study provides a novel framework for assessing emotional state in ALS, several limitations must be acknowledged. First, the small sample sizes in the “Muted” (N=5) and “Excessive” (N=4) subgroups limit the statistical power and generalizability of the findings. As a pilot study, these results should be viewed as a proof-of-concept rather than definitive clinical profiles. Second, although the clinician was blinded to the acoustic analysis, the “gold standard” for classification remains partially heuristic, relying on clinical intuition alongside patient feedback.

### Critical Evaluation of LLMs for Quantitative Emotional Assessment

The recent proliferation of Large Language Model (LLM) conversational agents for mental health support is remarkable [30]. However, our findings establish a crucial caveat: LLMs are currently unreliable for quantitative emotional assessment in this clinical population.

Our data demonstrates that LLMs give unreliable “diagnostics” when assessing a patient’s emotional state using the quantitative metrics from questions 1 to 4 of our protocol. The inability of LLMs to reproduce their own output (as indicated by uncertainty matching the score difference) raises a critical question: how can an LLM be trusted to deliver effective psychological support to an ALS patient if its initial assessment (or “diagnostic”) of their emotional state is fundamentally incorrect?

While we maintain this critical perspective, we also acknowledge that LLM performance is evolving rapidly, making future capabilities difficult to predict.

### Towards a Multi-Modal Assessment Approach and Clinical Limitations

Our core hypothesis is that a robust and reliable assessment requires a multi-modal approach. For reasons of scalability and practical ubiquity, our current work was restricted to the analysis of voice data exclusively. In ALS, speech analysis is particularly powerful as bulbar involvement and progressive motor degradation make voice an immediate, non-invasive proxy for underlying neurological function [31], [32]. We also note that the majority of patients in our cohort had spinal-onset ALS. Patients with bulbar-onset ALS were underrepresented because bulbar involvement more frequently impairs speech, and intact speech was required for participation in our study. This constitutes a limitation of the study and introduces a potential selection bias.

We recognize the inherent methodological limitation that any clinical discussion or formal collection of patient feedback may inherently affect or trigger the emotion being measured. To safeguard the objectivity of our analysis, the clinician was blinded to our conclusions until data collection was complete.

Furthermore, we confirmed established findings that a significant gap can exist between a patient’s subjective feedback and objective clinical findings (e.g., between breathing reporting and FVC percentage), meaning routine tests done in clinic may not offer a clear clue regarding a patient’s emotional management [33]. Similarly, standard sentiment analysis based on the VADER lexicon [15] was not sensitive enough to capture our classifications, suggesting that identifying ALS patient emotional management requires customized NLP methods trained on disease-specific language.

### Sex Differences in Emotional Management: Behavioral Driver

A notable finding was that Class 3 (Excessive Reaction) was populated mostly by men and Class 1 (Muted Reaction) mostly by women. This is counterintuitive given that the HADS score suggested a bias favorable to men, who typically report lower levels of internal anxiety (HADS scores) than women. This decoupling is highly significant: the classification is not a measure of general anxiety, but of how that anxiety is managed and expressed in the clinical setting, serving as a behavioral driver.

The average age of our cohort is close to 70 or older. We posit that the anxiety driving the Excessive Reaction in men is primarily functional anxiety related to the loss of traditional male roles of that generation and responsibilities concerning physical and financial function [34]. This ‘concern’ represents a specific, externalized form of distress over dependency and loss of control. This behavioral expression is validated by the clinical observation that these patients require significantly more clinician time, confirming that the perceived need for complex intervention is driven by the emotional presentation. This highlights the importance of using “concern”—a functionally oriented term—over “anxiety” in clinical protocols for ALS, as it appears to capture a critical, gender-influenced element of disease coping otherwise obscured by standard psychological screening tools.

We acknowledge that the sociocultural drivers of anxiety identified in this cohort (e.g., traditional male gender roles) may be specific to this age group (∼70 years) and geographic location, and may not translate to younger patients or different cultural settings.

### The Striking Decoupling: Emotion as the Driver, Dysarthria as the Filter

The psycho-acoustic analysis of speech revealed a striking, non-intuitive decoupling where the behavioral category may align well with the dominant dysarthria subtype. However, contrary to an interpretation that the classification is only a physical artifact, our findings confirmed by the time differences allocated to the patient by the clinician, suggest that emotion is the driver of the behavioral classification, while dysarthria may act as an acoustic filter or amplifier of that signal.

Because the psychoacoustic features (loudness, roughness, fluctuation) are the exact same features used to diagnose UMN vs. LMN damage, it is difficult to prove that the “Muted” group actually has an emotional state that is being masked. It remains possible that the classification is primarily measuring the physical severity of the dysarthria rather than a psychosocial coping mechanism. However, identifying properly dysarthria types in patient is so difficult for clinician [35] due in part by the mixed nature types often observed in patient, that clarifying the issue will require a far deeper study.

Overall, the features extracted from this classification appear as meaningful tools to support the clinical assessment.

Our classification tool and digital biomarkers propose a model where the patient’s observed emotional management strategy is not purely an acoustic artifact of dysarthria, but a confluence of psychological coping mechanisms filtered through the physical constraints of the dominant neuropathology. This demonstrates that these behavioral categories serve as readily observable non-invasive clinical proxies for both the dominant underlying neurological pathology and the patient’s need for psychosocial support.

## Conclusion: Noninvasive Assessment and Neurological Proxies

In this manuscript, we addressed the critical feasibility challenge of noninvasive assessment of emotional management in ALS patients by introducing a novel, functional classification system. This system identifies patients who exhibit a congruent, excessive, or muted reaction to their progressive disease, based on the proportionality between clinician-rated functional concern and self-reported ALS-FRS symptoms.

We demonstrated that while standard NLP tools, such as sentiment analysis, and current-generation LLMs provide unreliable quantitative diagnostics for this nuanced assessment, our novel classification method, relying primarily on voice analysis, offers a robust alternative. The method’s strength lies in its ubiquity, allowing seamless application across various clinical settings, including in-person sessions and telemedicine.

The most puzzling finding is the striking link between the behavioral classification and the patient’s underlying neuropathology. The observed emotional management strategy—driven by psychosocial coping mechanisms and requiring differential clinician time—could be superposed to the acoustic profile of Spastic Dysarthria for the muted group, and Flaccid Dysarthria for the excessive group.

This work provides a series of tools such as first plotting the patient evaluation in the graphic of figure 6 and second evaluating the speech acoustic features - capable of functioning in real-time to alert the clinical team to a patient’s specific emotional management profile and, indirectly, their potential bulbar phenotype. Such alerts can trigger a personalized and proactive strategy for disease management, psychosocial support, and targeted intervention. Looking forward, we plan first to integrate additional data channels from ubiquitous devices, such as oximeters and smartwatches, to further refine our multi-modal assessment, ultimately contributing to improved, personalized care for ALS patients. Second it will be very exciting to extend this feasibility study to a multi-center one that involved many more early staged ALS patients.

## Generative AI disclosure

In this study, Generative Artificial Intelligence (AI) was employed for both methodological development and comparative analysis. Specifically, ChatGPT (OpenAI) was used to assist in the manual extraction and development of a key term dictionary for Natural Language Processing (NLP) topic classification. Additionally, current-generation large language models (LLMs), including ChatGPT, Gemini (Google), and Claude (Anthropic), were utilized to evaluate their ability to quantitatively score patient anxiety levels based on clinical transcripts. These LLM-generated scores were compared against established clinician ratings to assess the models’ reproducibility and reliability for clinical emotional assessment. The researchers confirm that while these tools assisted in specific data classification and analysis tasks, they did not replace expert clinical supervision or the primary data-driven findings of the study.

## Ethics statements

### Studies involving animal subjects

No animal studies are presented in this manuscript.

### Studies involving human subjects

No human studies are presented in the manuscript.

No potentially identifiable images or data are presented in this study.

## Data availability statement

The original contributions presented in the study are included in the article/supplementary material, further inquiries can be directed to the corresponding author/s.

## Figures

### Annex Figure

**Figure A1.**
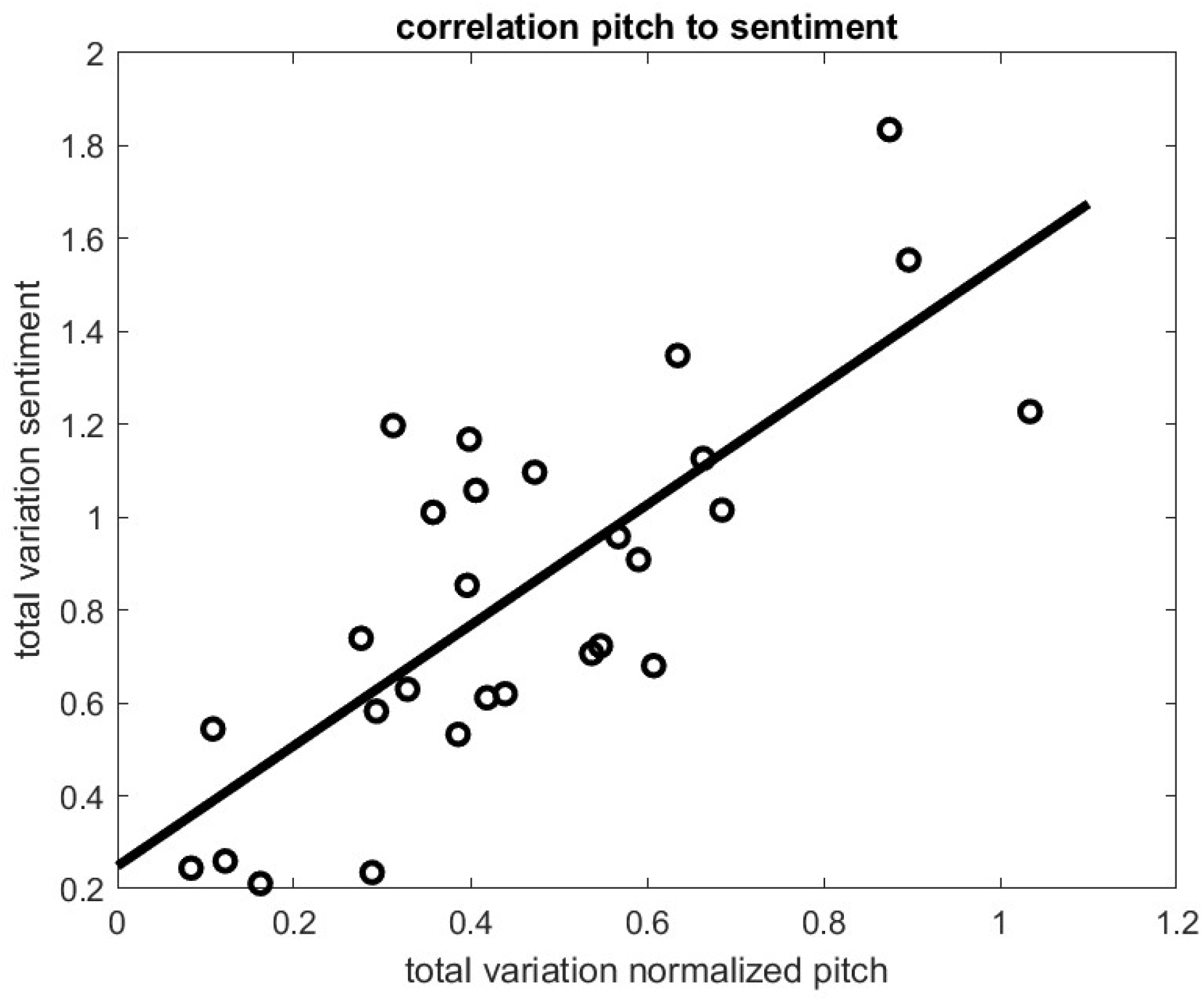
Total Variation of Sentiment versus Total Variation of Pitch during the clinical examination; correlation coefficient is about 0.7 and p value is 10^-6. The total variation of a real-valued function $f$ defined on a closed interval $[a, b]$ is a measure of the “total oscillation” or “roughness” of the function over that interval.

**Figure A2.**
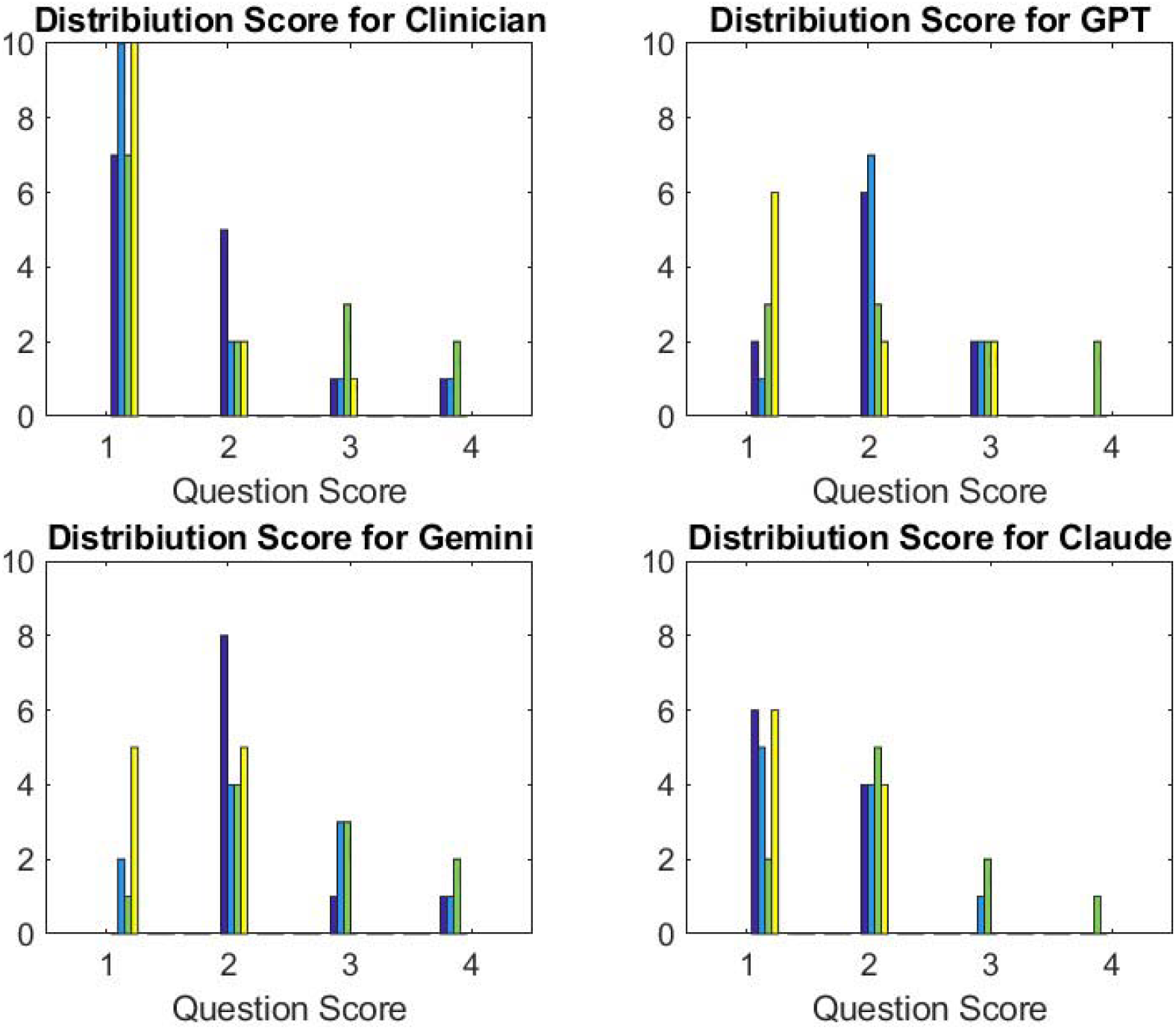
Comparison of the Distribution of the 4 Question Scores between Clinician (top left) and LLM with respectively Chat GPT (top right), Gemini (bottom left) and Claude (bottom right). We observe a larger distribution of LLM score than for the clinical one: LLM score are usually higher than clinical one, Claude being the closest to the clinical score.

### Annex LLM

#### LLM Prompt with DATA SET 0

From the following conversation, can you score successively from 0 to 3 the anxiety of the patient related to 1. breathing and Mobility issues, 2 Nutrition issues, 3 Finance issues. 0 meaning no anxiety, 1 meaning mild anxiety, 2 meaning moderate anxiety and 3 meaning extreme anxiety:

#### LLM Prompt with DATA SET 1

From the following conversation, can you score successively from 0 to 3 the anxiety of the patient related to 1. breathing issues, 2 Mobility issues, 3 Nutrition issues, 4 Finance issues. 0 meaning no anxiety, 1 meaning mild anxiety, 2 meaning moderate anxiety and 3 meaning extreme anxiety:

